# Cross-sectional evolution of pain management policies and practices in Portuguese pediatric emergency departments

**DOI:** 10.1101/2022.07.06.22277222

**Authors:** André Garrido, Ana Rute Manuel, Ricardo Araújo, Inês Mascarenhas, Helena Almeida, Clara Abadesso

**Author notes:** Address correspondence to: André Garrido; Departamento de Pediatria, Hospital Prof. Doutor Fernando Fonseca, E.P.E., IC19, 2720-276 Amadora.

## Abstract

The prevalence of pediatric pain, either related to the child’s hospital visit or as a result of diagnostic and/or therapeutical interventions, is of primordial importance in pediatric emergency departments (PEDs).

In this study, we evaluate the evolution of pain assessment and management in Portuguese PEDs through eleven years. To this end, we prepared a questionnaire addressed to head physicians of 45 the Portuguese PEDs in 2007 and compared the responses to those provided in 2018, where we also posed these questions to nurse managers.

Pain assessment in Portuguese PEDs has significantly improved, namely with establishment of local protocols and widespread use of pain scales. However, effective adoption of pain management remains insufficient, as mild to moderate pain is still far from being universally treated. Nonetheless, there seems to be an adequate treatment of severe pain and respective common use of opioids, but correct practices were not generally adopted when specific types of pain were analyzed. Procedural sedation and use of non-pharmacological techniques has significantly increased, but are not yet universally practiced. These inadequacies are reflected by the staff’s perception that pain management remains suboptimal, and more training is needed, effectively urging for a nationwide plan and better knowledge translation of correct pediatric pain management.

## INTRODUCTION

In recent decades, the recognition of pain relief as a fundamental right resulted in significant progress in understanding and assessing pediatric pain. Indeed, children experience pain distinctively from adults, as they exhibit differences in the neurobiology of pain(Pancekauskaite and Jankauskaite, 2018). Pain is a chief complaint in the Pediatric Emergency Department (PED), being prevalent in more than 60% of the cases at admission(Marzona et al., 2019; Taylor et al., 2008). In this context, pain must be promptly assessed and treated, regardless the underlying condition. Despite ongoing improvements in pain relief, pediatric pain management is still insufficient worldwide(Ali et al., 2014; Farhat et al., 2013; Ferrante et al., 2013; Herd et al., 2009; MacLean et al., 2007; Williams et al., 2019). Ironically, pain induced by health professionals during various routine procedures is often underestimated, being amongst the most common sources of acute pain at the PED(Pancekauskaite and Jankauskaite, 2018; Senger et al., 2021).

Pain treatment must be central to pediatric action as any form of oligoanalgesia has short- and long-term consequences. In the short-term, acute pain amplifies pain perception, anxiety and fear, which prevents accurate physical examination and delays necessary interventions. Consequently, potential needless exacerbation of the child’s condition and the increased length of stay at the PED could be avoided. In the long-term, untreated pain may affect a child’s emotional well-being and increase the possibility of chronic pain(Anand and Scalzo, 2000; Noel et al., 2017; Taddio et al., 2009; Williams et al., 2019; Young, 2005). It is, thus, crucial that pain is accurately assessed and nurse-initiated pain treatment protocols are instituted. Pain treatment must be prioritized and optimized, using non-pharmacological interventions to reduce distress and anxiety, combined with pharmacological treatment, proportional to pain severity(Bailey and Trottier, 2016; Bauman and McManus, 2005; Drendel and Ali, 2017; Fein et al., 2012; American Academy of Pediatrics, 2001; Ruest and Anderson, 2016).

Allied to this avoidable scenario is the fact that pediatric pain has only started to receive attention in the 1980s(Schechter, 2008). Portugal established the National Day Against Pain as early as 1999 and created a National Plan Against Pain in 2001(Diniz et al., 2001) which has been periodically updated(Direção-Geral da Saúde, 2017). In 2003, pain was considered the “5^th^ vital sign”, which enforced systematic records of pain intensity(Direção-Geral da Saúde, 2003) and, in 2010, pediatric-specific guidelines were published by the Ministry of Health(Direção-Geral da Saúde, 2010). Since then, specific pediatric guidelines were published covering pain management in neonates, children with cancer, and invasive procedures(Direção-Geral da Saúde, 2012a-c). Despite this favorable context, no studies demonstrate to what degree these recommendations have been converted into local protocols at the Portuguese PEDs. Also, there is no national data regarding the evolution of pain control over the years, and how it compares with other countries.

This study aims to describe the policies and reported practices of pain management at a national level by comparing the results between two surveys performed in 2007 and 2018 in a sample of Portuguese PEDs.

## METHODS

### Study Design

In 2007, a structured questionnaire was mailed to all 45 Portuguese hospitals with PEDs, to be answered by the head physician. A similar questionnaire designed in Google® Forms was emailed to the same PEDs in 2018 (Supplemental Material), to be answered by each responsible physician and nurse. Questions included are discriminated in the results section. Most questions had a Likert scale: never, <50% of times; > 50% of times, always. Responses to questionnaires were collected along 2007 and between the 1^st^ of August of 2018 and the 31^st^ of July of 2019, respectively. We excluded all repeated questionnaires, and we only accepted the original responses when questionnaires were repeated. Most of the questions required an answer to move forward in the 2018 questionnaire. Whenever the 2007 questions remained unanswered, we adjusted the sample accordingly.

### Statistical Analysis

All statistical analyses were performed using XLSTAT 2018.1.1, using a 95% confidence interval for all statistics. All P-values were obtained by the asymptotic method and are two-sided. We assumed for all statistics the independence of answers between hospitals and equal variances. We have four types of samples (see Fig. 1):

**Figure 1.**
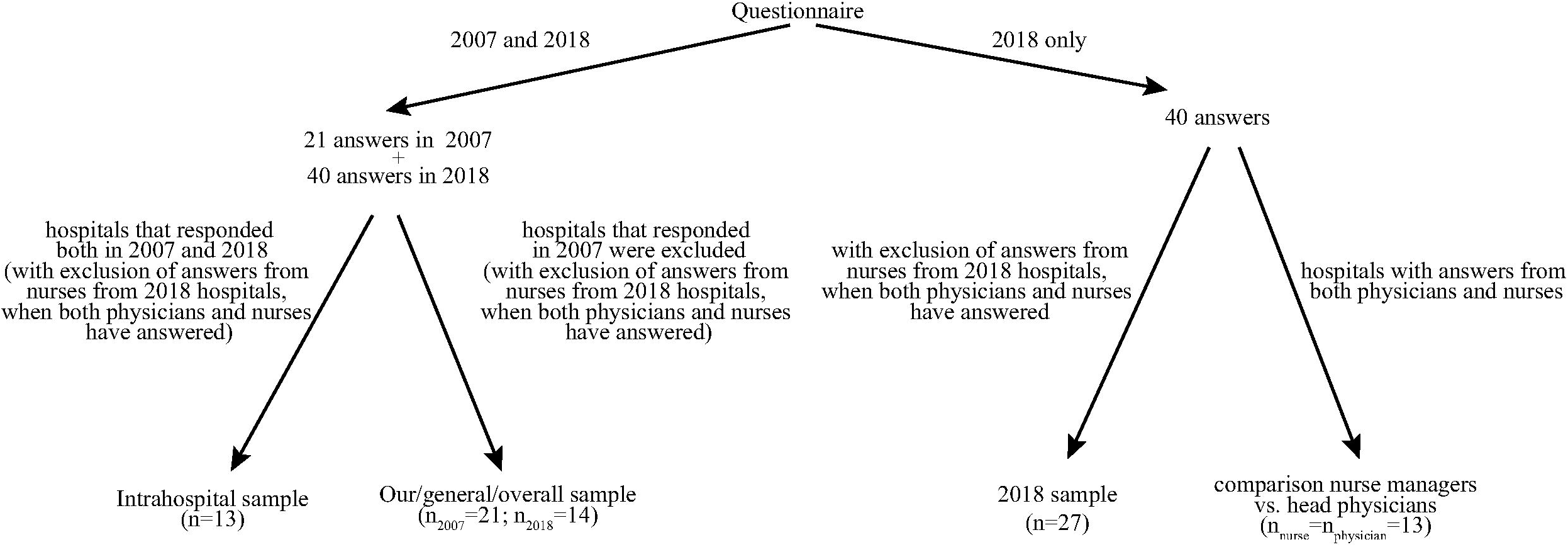
Rationale for the various analyses performed and corresponding sample adaptations.

1. our general sample of Portuguese hospitals (hereafter designated as “general sample” or “our sample”) is the main focus of our study; we assume to be a representative sample of the general scenario in the Portuguese PEDs as it compares the responses from 2007 and 2018 eliminating repeated hospitals for 2018 that also provided replies to the questionnaires in 2007; we used a Student’s t-test (n_2007_=21; n_2018_=14) for these analyses.
2. the intrahospitalar time series aims to look at the consistency of practices within the same hospitals in the two sampled time points (2007 and 2018); for these comparisons we used a paired t-test (n=13).
3. the 2018 only sample analyses responses to questions only present in the 2018 questionnaire; we excluded repeated answers from the same hospital, excluding answers by nurse managers in these cases (n=27);
4. comparison between head physician and nurse manager responses evaluates if physicians and nurses responded differently in 2018; we used only the responses from hospitals that had both responded and we analyzed the data using a paired t-test (n_nurses_=n_physicians_=13).

When answers were originally formatted as “never”, “<50% of times”, “>50% of times” and “always” we statistically looked at the tendency of the results and, for this reason, the intermediate answers cannot be quantitatively objectified.

## RESULTS

### Sample

In 2007, there were 21 different hospital answers to the questionnaire, all from the head physicians. In 2018, there was a total of 40 answers, of which 23 were made by the head physician and 17 by the nurse manager. However, 13 of them were provided by both the head physician and the nurse manager from the same hospital. This means that 47% of the 45 portugueses PEDs responded in 2007 and 60% in 2018. Table 1 shows the characterization of the responses as well as the medical specialties operating in different PEDs. The annual emergency episodes reported was on average ∼31910 cases (SD=11286.9) in 2007 and 40943 cases (SD=14737.9) in 2018.

**Table 1.**
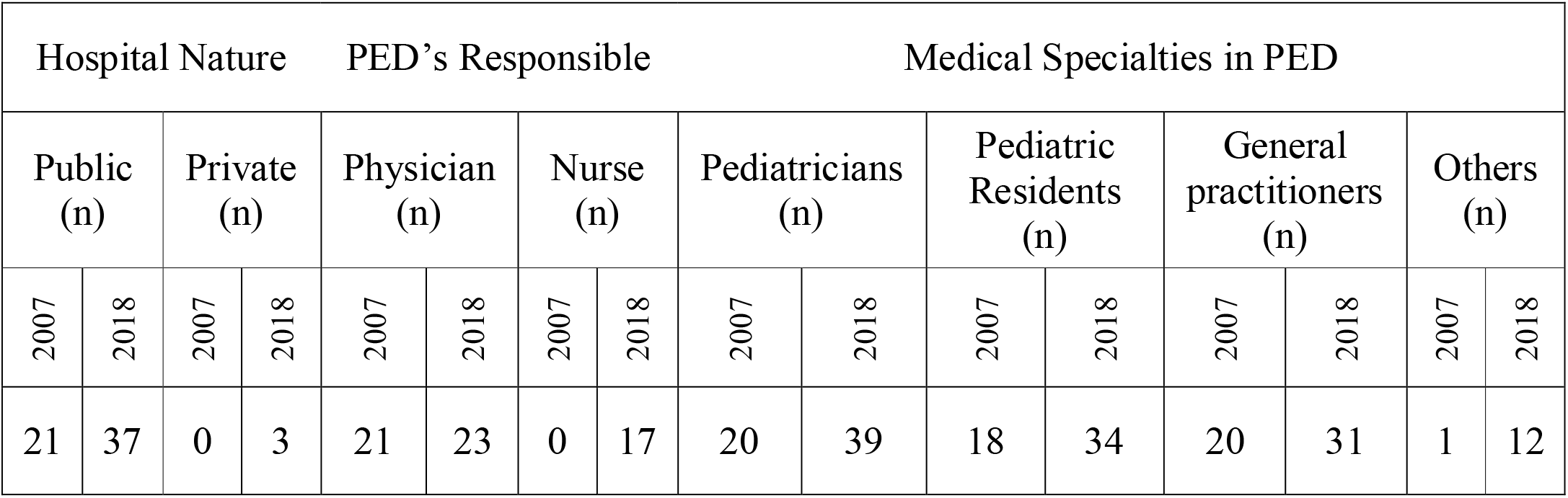
Characterization of the responses to questionnaires from Portuguese PEDs responsible staff in 2007 and 2018; PED – Pediatric emergency department.

### Pain Protocol

#### Prevalence of protocols for pain management

There is a statistically significant increase in the number of hospitals adopting a protocol for pain management, from 52% of hospitals in 2007 to 93% in 2018 (P=0.01). However, 15% of hospitals reported differently their existence between nurse managers and head physicians. Among the 25 hospitals in 2018 that reported adopting a protocol, these were introduced on average 8 years before the questionnaire (SD=5.2 years).

Among our sample of 27 hospitals for 2018, 81% affirmed to do analgesia in triage by protocol (no different responses were observed between nurses and physicians from the same hospitals).

### Pain Assessment

#### Prevalence of pain assessment

Among our general Portuguese hospitals sample, there is a statistically significant increase in the number of times pain is assessed reported by the responsible staff (P=0.006). Whereas in 2007 there were 43% of hospitals reporting assessment of pain in at least 50% of emergency incidents, in 2018 all hospitals did so. However, when we consider hospitals that assess pain in all PED episodes, there is a non-significant increase from 38% to 64% (P=0.14).

#### Pain assessment: who, method, pain scales, and locus

In our sample, when comparing 2007 to 2018, there was a statistically significant increase of nurses providing pain assessment, from 52% to 100% (P=0.001). However, the contribution that physicians provide for pain assessment was maintained between the years studied, 57% in 2007 and 50% in 2018 (P=0.69). Notably, no hospitals considered parent’s assessment of pain in 2018, while in 2007 nearly 50% percent of the hospitals did (P<0.001).

In our sample, hospitals started adopting significantly more pain scales from 2007 to 2018, 62% versus 100% (P=0.01). The pain scales used the most, by decreasing order reported in 2018, were: Numeric (n=30); Faces (n=23); Face, Legs, Activity, Cry, Consolability (FLACC) (n=20), Analogic (n=11), Neonatal Infant Pain Scale (NIPS) (n=9), Portuguese Triage Group Scale (4), Faces Pain Scale – Revised (FPS-R) (n=3), Face, Legs, Activity, Cry, Consolability – Revised (FLACC-R) (n=2), Hétero Evaluation Douleur Enfant (HEDEN) and Échelle Douleur Inconfort Nouveau-Né (EDIN) scales (both n=1).

### Analgesia for Different Types and Presentations of Pain

The characterization of how pain is treated for different types of pains can be found in Table 2.

**Table 2.**
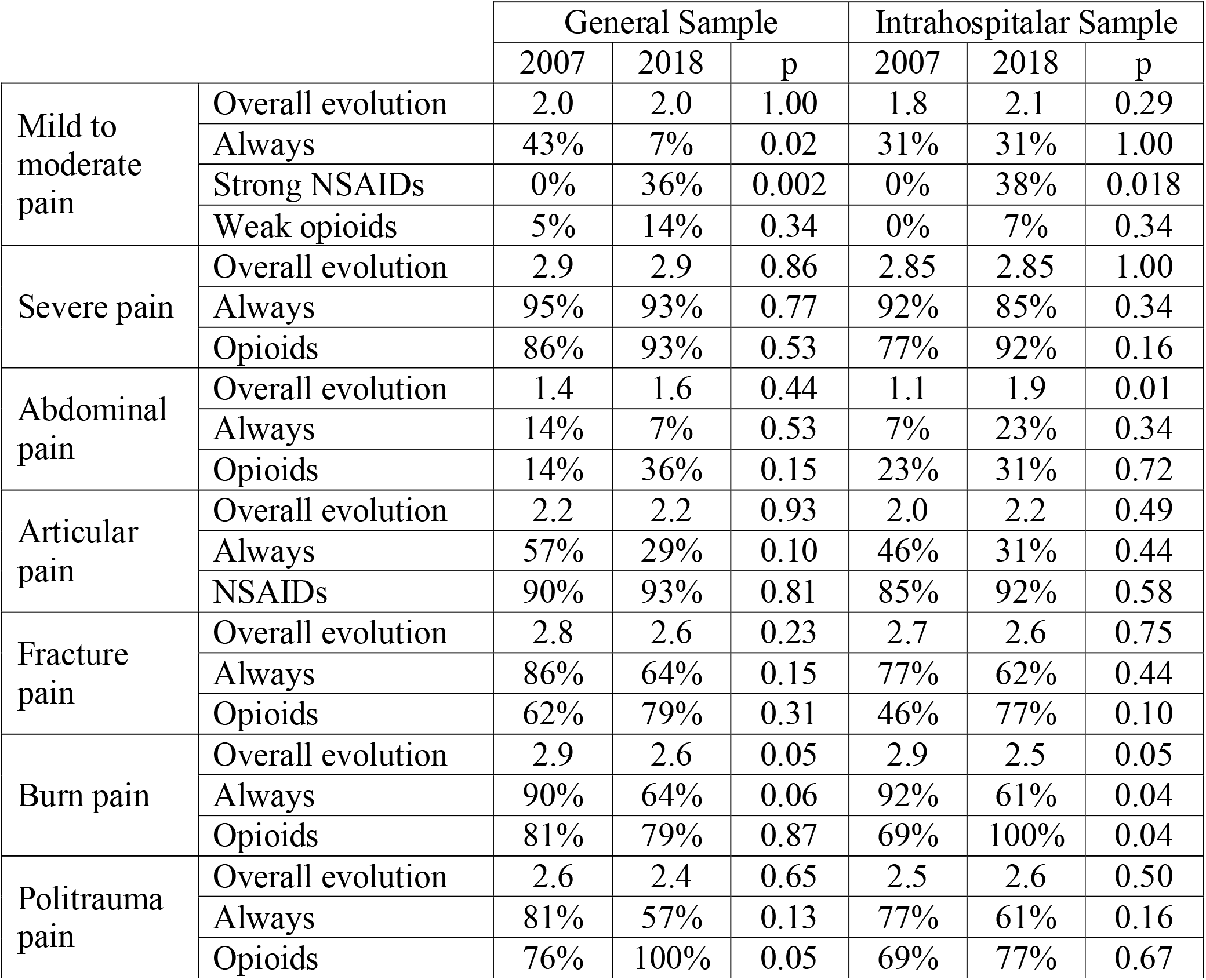
Characterization of pain’s treatment for different types of pains concerning the general and the interhospitalar samples; overall evolution - the mean of a scale from 0 to 3, in which 0 means never treated, 1 means <50%of times, 2 means >50% of times and 3 means always treated; always – concerns to the percentage of hospitals in which pain is always treated; strong NSAIDs / weak opioids / opioids – concerns to the percentage of hospitals in which these types of analgesics are used for pain’s treatment.

#### Analgesia of mild-moderate and severe pain

Concerning the use of analgesia for mild to moderate pain results were similar in 2007 and 2018. Always using analgesia in this situation had a significant decrease from 43% to 7% in our sample (P=0.02). There is a very significant increase of use of strong nonsteroidal anti-inflammatory drug (NSAIDs) like diclofenac, ketorolac and metamizole from 2007 to 2018 for this type of pain across our sample (P=0.002) and intrahospitally (P=0.018). None of the hospitals used strong NSAIDs in 2007, whereas in 2018, 36% were already using them.

There is a widespread use of analgesia for severe pain both in 2007 and 2018, as also the use of opioids in these situations.

#### Analgesia for abdominal, articular, fracture, burn, and politrauma pains

For abdominal pain there was a non-statistically significant increase for analgesics use in our sample but a significant increase intrahospitally. For the use of analgesics for articular pain treatment, there was no evolution between the sampled years. There was a non-statistically significant reduction of use of analgesics for fracture pain. We registered one hospital that only used paracetamol for the treatment of fracture pain, both in 2007 and 2018. There was a statistically significant decrease in use of analgesics for burn treatment in our sample and a near non-statistically significant in the intrahospitalar sample. There was no statistically significant variation of analgesics use in our sample of Portuguese hospitals for politrauma treatment.

### Procedural Analgesia and Sedation

Characterization of different procedures sedoanalgesia can be found in Table 3.

**Table 3.** Characterization of pain’s treatment of different procedures concerning the general and the interhospitalar samples; overall evolution - the mean of a scale from 0 to 3, in which 0 means never treated, 1 means <50%of times, 2 means >50% of times and 3 means always treated (except for local analgesia for intramuscular injections which was closed question, answering yes/no); never / always – concerns to the percentage of hospitals in which pain is never and always treated, respectively.

|  |  | General Sample |  |  | Intrahospitalar Sample |  |  |
| --- | --- | --- | --- | --- | --- | --- | --- |
|  |  | 2007 | 2018 | p | 2007 | 2018 | p |
| Venoanalgesia | Overall evolution | 0.9 | 1.2 | 0.37 | 0.9 | 1.1 | 0.66 |
|  | Never | 43% | 13% | 0.06 | 46% | 23% | 0.19 |
|  | Always | 14% | 14% | 1.00 | 15% | 8% | 0.58 |
| Local analgesia for intramuscular injections | Overall evolution | 62% | 50% | 0.50 | 75% | 50% | 0.22 |
| Analgesia for lumbar puncture | Overall evolution | 1.0 | 1.1 | 0.52 | 0.9 | 1.1 | 0.08 |
|  | Never | 24% | 7% | 0.21 | 23% | 8% | 0.16 |
| Sedation for lumbar puncture | Overall evolution | 0.3 | 0.9 | 0.006 | 0.4 | 0.7 | 0.16 |
|  | Never | 67% | 21% | 0.008 | 61% | 46% | 0.34 |
| Analgesia for fracture reduction | Overall evolution | 1.1 | 1.6 | 0.14 | 1.0 | 1.5 | 0.05 |
|  | Never | 33% | 7% | 0.07 | 38% | 8% | 0.04 |
|  | Opioids | 10% | 43% | 0.02 | 7% | 7% | 1.00 |
| Sedation for fracture reduction | Overall evolution | 0.5 | 1.4 | 0.001 | 0.3 | 1.1 | 0.009 |
|  | Never | 61% | 0% | <0.001 | 69% | 15% | 0.012 |
| Analgesia for wound disinfection and suture | Overall evolution | 1.5 | 1.4 | 0.85 | 1.4 | 1.2 | 0.58 |
|  | Never | 19% | 0% | 0.09 | 23% | 0% | 0.08 |
| Sedation for wound disinfection and suture | Overall evolution | 0.3 | 0.8 | 0.02 | 0.3 | 1.0 | 0.02 |
|  | Never | 71% | 21% | 0.003 | 69% | 23% | 0.03 |

#### Analgesia for venopuncture and use of lidocaine for intramuscular injections

For venopuncture, there was an increase in use of topic analgesia from 2007 to 2018 in both samples, but such results were not statistically significant.

There was no statistically significant change in the use of local lidocaine before intramuscular injections in our sample, 62% in 2007 and 50% in 2018 (P=0.50). However, in 2018, there is a statistically significant difference between the answers within the same hospital provided by physicians and nurses with 46% physicians confirming the use of lidocaine, whereas 85% nurses reported the use of this technique (P=0.02).

#### Analgesia and sedation for lumbar puncture

Although there was an increase in our sample and intrahospitally for the use of analgesics for lumbar punctures, namely topic analgesia, it was not statistically significant. Concerning the use of sedation for the lumbar puncture procedure, there was a statistically significant increase for our sample.

#### Analgesia and sedation for fracture reduction

The increase of analgesics use for fracture reduction was not statistically significant within our sample, but near-significant intrahospitally. There was a significant increase in use of sedation for this procedure in the general panorama and intrahospitally. Fracture reduction procedures were performed within PEDs in 48% of the 27 hospitals interviewed in 2018.

#### Analgesia and sedation for wound suture

Concerning the use of analgesia for wound disinfection and suture, is similar in both periods. However, there is a statistically significant increase in use of sedation for this procedure. Among 27 hospitals interviewed in 2018, 67% performed the wound suturing and disinfection procedure in the PED.

#### Use of sucrose, lidocaine/prilocaine patches (EMLA®), and equimolar mixture of oxygen and nitrous oxide (EMONO)

There was a statistically significant increase in terms of the use of sucrose for infants <6 months years old for painful procedures in our sample from 52% of hospitals in 2007 to 100% in 2018 (P=0.001).

Among 27 hospitals interviewed in 2018, 85% use EMLA® and 37% had EMONO.

#### Medical responsibility for IV procedural sedoanalgesia and pre-procedural risk evaluation

In 2018, intensivists or anesthetists supported the sedation prescription in 85% of hospitals and this prescription was performed exclusively by these physicians in still 7% of hospitals. The pre-sedation risk evaluation is performed in 93% of hospitals and 89% of hospitals registered this risk and the sedation procedure.

### Pain reassessment after analgesia

Concerning if efficacy of pain treatment is assessed, the results were already high in 2007, but there was a slight increase across the general sample, 81% in 2007 to 93% in 2018, but not statistically significant (P=0.34).

### Non-Pharmacological Interventions

The increase in the use of non-pharmacological interventions in our sample from 52% in 2007 to 79% in 2018 was not statistically significant (P=0.12). However, it is statistically significant when such practices are compared intrahospitally, from 50% in 2007 to 92% in 2018 (P=0.02). In 2018, there is no statistically significant difference between the answers within the same hospital provided by physicians and nurses concerning the use of these techniques (P=0.58).

Among 27 hospitals interviewed in 2018, the parents are always or most of the times present during procedures in 93% of hospitals (Table 4). However, only 51% of hospitals confirmed using the parent’s lap always or most of the times. In 2018, 26% of hospitals reported using forced immobilization most of the times. Interestingly, there is a statistically significant difference between the answers within the same hospital provided by physicians and nurses, with physicians reporting the use of force more often than nurses (P=0.04).

**Table 4.** Characterization of the percentage of hospitals in which parental presence, parent’s lap and forced immobilization are used in 2018.

|  | Always | >50% of the times | <50% of the times | Never |
| --- | --- | --- | --- | --- |
| Parental presence | 37% | 56% | 7% | 0% |
| Parent's lap | 7% | 44% | 30% | 18% |
| Forced immobilization | 0% | 26% | 70% | 4% |

### General Use of Opioids

The general use of opioids did not change significantly both across the general and the intrahospitalar samples (P=0.76 and P=0.67, respectively). Similarly, no significant variations were observed when weak and strong opioids were compared alone. Weak opioids use decreased from 48% in 2007 to 36% of hospitals in 2018 (P=0.50). Strong opioids had a marginal increase from 61% to 64% of hospitals (P=0.59). In 2018, opioids were prescribed exclusively by intensivists or anesthetists in still 7% of hospitals.

### Training and Adequacy of Pain Treatment

#### Staff’s perception of pain treatment adequacy and need for training

Concerning the perception of adequacy of pain treatment practiced in the sampled Portuguese hospitals, there seems to be a slight decrease in our general sample from 67% in 2007 to 57% in 2018 (P=0.58), but a slight increase intrahospitally from 58% in 2007 to 67% in 2018 (P=0.67). However, none of these results are statistically significant (P=0.58 and P=0.67, respectively). At the same time, there seems to be an increased will for additional training when our sample is considered, from 86% in 2007 to 100% in 2018 but this result was not statistically significant too (P=0.15).

#### Staff’s perception for main reasons of pain treatment inadequacy

The main reasons appointed for an inadequacy of pain treatment in 2007 were, from the most to the less chosen, fear of hiding symptoms or signs (38% of hospitals), difficulty of a correct pain assessment (33%) and fear of opioid side effects (24%). In 2018, the most chosen reasons were fear of hiding symptoms or signs (50% of hospitals), lack of time (33%), fear of opioid side effects (29%) and medical inexperience in prescribing analgesics (29%).

## DISCUSSION

From 2007 to 2018, despite significant improvements in pain assessment in Portuguese PEDs, the routine practice of pain treatment protocols had not ensued. Mild to moderate pain remains infrequently treated, although severe pain treatment and corresponding use of opioids is prevalent. Similarly, although the use of sedation increased significantly for many painful procedures, analgesia during these procedures persists being scarce. In line with these results, there is still a considerable proportion of hospitals that acknowledge inadequate treatment of pain. Fear of children hiding symptoms/signs and hesitancy concerning opioid use account for the most prevailing reasons for unsatisfactory treatment, mirroring previously reported results(Craig et al., 1996; Czarnecki et al., 2019; McGrath and Frager, 1996; American Academy of Pediatrics, 2001). In the other direction, the use of non-pharmacological techniques for pain control has improved, with multiple hospitals reporting the child sitting on the caregiver’s lap during painful procedures, but others, however, still frequently use forced immobilization, a practice that is against ethical and legal considerations in health care(Bailey and Trottier, 2016; Direção-Geral da Saúde, 2012a; Ruest and Anderson, 2016; Trottier et al., 2019; Wente, 2013). Nevertheless, the widespread desire of further training on the treatment of pain by health practitioners offers a sign of hope for the future.

The implementation of local protocols for pain management and use of recommended pain scales is now a highly prevalent practice in Portugal (93% and 100%, respectively), reflecting the Portuguese and international guidelines and practices(Direção-Geral da Saúde, 2010; Fein et al., 2012; Krug et al., 2009; Marzona et al., 2019; Schug et al., 2020; Williams et al., 2019; Young, 2017). With the universal use of pain scales, nurses settled as the main pain assessors and parent’s contribution has significantly fallen. In line with the evolution in Italy(Benini et al., 2020), there was a significant increment in pain assessment in Portugal from 2007 to 2018, but not yet universally adopted as suggested(Fein et al., 2012; Marzona et al., 2019). North-American PEDs have universally implemented the use of pain assessment protocols (Haupt et al., 2021), which should have downstream effects leading to more adequate and personalized treatments, such as subsequent administration of analgesia.

In fact, analgesia in triage by protocol was not yet a generalized practice by 2018 in Portuguese PEDs (81%), performing between Italian (68%) and Australian (92%) PEDs(Benini et al., 2020; Herd et al., 2009). Similarly, mild to moderate pain analgesia is not administered during a significant proportion of episodes in Portuguese PEDs and the 11-year evolution was significantly unfavorable when considering the PEDs that always do it. This is a worrying scenario as children often receive non-adequate pain relief treatment(Brudvik et al., 2017). However, there was a significant improvement in administration of strong NSAIDs for this type of pain (36%). Opioids are practically unused in this setting, despite the broad consensus for its use for moderate pain in lower doses(Bailey and Trottier, 2016; Bauman and McManus, 2005; Fein et al., 2012; Ruest and Anderson, 2016; Young, 2017).

Unlike previous studies(Brudvik et al., 2017; Senger et al., 2021), there was high percentage of treatment of severe pain for the years studied, as well as widespread use of opioids in this context.

Concerning some specific pain treatments, the 11-year evolution has not been generally satisfactory. For instance, abdominal and articular pains remain infrequently treated with few hospitals reporting treatment in all instances (22% on average), similar to results for other countries(Farhat et al., 2013; Kleiber et al., 2011). Fracture-, burn- and politrauma-associated pains are reported to always be treated on average in 62% of hospitals. If we consider that these situations are usually associated with moderate-severe pain, this percentage is not consistent with the general answer about treatment of severe pain. Alongside, the generalized use of opioids has been reported for politrauma associated pain (100%), but less so for fracture- and burn-associated pains (79% for both).

Beyond sparse adherence to specific pain treatment with analgesics, the frequency of analgesia for various medical procedures remains uncommon, and incorrect practices continue. Notably, however, there is a slight reduction of hospitals that never use analgesics for these procedures, but analgesia remains rarely used for venipuncture, below the average in Europe and Canada(Ali et al., 2014; Sahyoun et al., 2021), but in line with other international reports(Ali et al., 2014; Kleiber et al., 2011). National and international recommendations, however, clearly suggest the implementation of analgesia when performing venipuncture(Bailey and Trottier, 2016; Bauman and McManus, 2005; Direção-Geral da Saúde, 2012a; Fein et al., 2012; Ruest and Anderson, 2016; Trottier et al., 2019). On the other hand, sucrose is now widely used in Portuguese PEDs, contrasting with other nations where this practice is less frequently used(Kleiber et al., 2011). Fortunately, sedation for painful procedures incremented considerably and, in fact, few hospitals never used sedation during these practices, in spite of EMONO unavailability in most Portuguese PEDs. Indeed, the availability of EMONO is below the European mean(Sahyoun et al., 2021). In a few Portuguese PEDs the wrong practice of limiting opioid and sedation prescription to intensivists/anesthetists persists (7% for both)(Coté and Wilson, 2019; Cravero, 2009; Cravero et al., 2006; Fein et al., 2012). This reflects the reduced specific training of general pediatricians on procedural analgesia and sedation.

Our study has several limitations including (1) the absence of control for non-response biases, i.e., the hospitals that responded to questionnaires probably had more interest in this subject; (2) the responses of the head physicians and nurse managers can be biased by different emphasis in the course of their education; (3) the sample size despite representing 60% of Portuguese PEDs is still statistically small, some results would have improved statistical power given larger samples; (4) non-pharmacological techniques were explored less objectively than pharmacological techniques, limiting the evaluation of the desired multimodal approach to pediatric pain(American Academy of Pediatrics, 2001). Despite these limitations, our cross-sectional study offers an insightful view over a decade of pain treatment and assessment at a countrywide level, allowing the definition of a clear roadmap to improve the quality of treatment in Portuguese PEDs.

In conclusion, Portugal is in line with other countries concerning the assessment and treatment of pain, but erroneous and inadequate procedures are still common. Fortunately, the creation of local pain protocols has significantly expanded, which contributed to a significant increment in pain assessment and to the widespread adoption of pain scales. Also, the treatment of severe pain remained satisfactory and corresponding use of opioids, but there is room for improvement as this is not a universal practice yet. Sedation use has also significantly increased in many painful procedures, but analgesia continues relatively uncommon. Though, the treatment of mild to moderate pain is far from being a generalized practice. A nationwide, articulated action is urgent. There are several educational resources available in Portugal released by the Directorate-General of Health(Direção-Geral da Saúde, 2010; Direção-Geral da Saúde, 2012a-c) and the Portuguese Association for the Study of Pain (available at https://www.aped-dor.org/). Knowledge translation is of outmost importance. The widespread of the already existing resources, as well as promoting local training specific for pain management and procedural analgesia and sedation in PED is crucial. Additionally, further scientific research including: (1) evaluation of efficacy of newly adopted strategies, (2) objective periodic evaluations of the healthcare system and, also (3) transnational collaborations at an Iberian or pan-European scale to promote pain research and allowing studies with large sample sizes are crucially necessary.

## Supporting information

Questionnaire

## Data Availability

All data produced in the present work are contained in the manuscript

## Acknowledgements

We are grateful to all nurses and physicians that kindly answered our questionnaire.

## Funding Acknowledgements

This research received no specific grant from any funding agency in the public, commercial, or not-for-profit sectors.

