## Supplementary material for "Cross-sectional evolution of pain management policies and practices in Portuguese pediatric emergency departments": Questionnaire

Questionnaire – Management of pediatric pain in the pediatric emergency department

Dear colleague,

We ask for 10 minutes of your time to answer this questionnaire.

We have the goal of exploring the actual practices of pain management in national pediatric emergency departments. To do so, I send you this questionnaire which I would like you to respond accordingly.

This questionnaire was already sent in 2007 and the results presented in March of the same year at the second meeting of the pediatric department of Hospital Prof. Dr. Fernando Fonseca.

Your participation is totally voluntary and without any kind of prejudice if you decide to not participate. The information shared will be protected and exclusively accessed by the team from the study. Every professionals of this team are restricted by professional secrecy.

Thank you for your time. Please, if you have any questions or need any additional information, feel free to contact us:

André Garrido – Pediatric Resident –
Clara Abadesso – Pediatrician – 

Email

____________________________________________

What is your role in the pediatric emergency department (PED)?

☐Physician – the head physician

☐Nurse – the nurse manager

Which is your hospital?

___________________________________________

Hospital type:

☐General hospital

☐Pediatric hospital

Average number of PED visits:

_______________________

Which type of physicians work at your PED?

☐Pediatricians

☐Pediatric residents

☐General practitioner

☐Others

Pain Protocol and Assessment

Is there a protocol for pain management at your PED?

☐Yes

☐No

If yes, for how long?

______________________

Is analgesia done in triage?

☐Yes

☐No

How frequently is pain assessed at your PED?

☐Always

☐> 50% of the times

☐< 50% of the times

☐Never

Who assesses pain?

☐The nurse from triage

☐The physician

How is it done?

☐Nurse’s impression

☐Physician’s impression

☐Parent’s impression

☐Pain assessment scale

☐Other

Which pain assessment scales are used?

______________________________________________________________________

Is pain reassessed after analgesia?

☐Yes

☐No

Analgesia for Different Types and Presentations of Pain

How frequently is analgesia done for mild to moderate pain?

☐Always

☐> 50% of the times

☐< 50% of the times

☐Never

Which drugs are used to do it?

☐Paracetamol

☐Ibuprofen

☐Diclofenac

☐Ketorolac

☐Dipyrone

☐Other NSAIDs

☐Tramadol

☐Morphine

☐Fentanyl

☐Other opioids

☐Other

How frequently is analgesia done for severe pain?

☐Always

☐> 50% of the times

☐< 50% of the times

☐Never

Which drugs are used to do it?

☐Paracetamol

☐Ibuprofen

☐Diclofenac

☐Ketorolac

☐Dipyrone

☐Other NSAIDs

☐Tramadol

☐Morphine

☐Fentanyl

☐Other opioids

☐Other

How frequently is analgesia done for abdominal pain?

☐Always

☐> 50% of the times

☐< 50% of the times

☐Never

Which drugs are used?

☐Paracetamol

☐Ibuprofen

☐Diclofenac

☐Ketorolac

☐Dipyrone

☐Other NSAIDs

☐Tramadol

☐Morphine

☐Fentanyl

☐Other opioids

☐Other

How frequently is analgesia done for articular pain?

☐Always

☐> 50% of the times

☐< 50% of the times

☐Never

Which drugs are used?

☐Paracetamol

☐Ibuprofen

☐Diclofenac

☐Ketorolac

☐Dipyrone

☐Other NSAIDs

☐Tramadol

☐Morphine

☐Fentanyl

☐Other opioids

☐Other

How frequently is analgesia done for bone fracture associated pain?

☐Always

☐> 50% of the times

☐< 50% of the times

☐Never

Which drugs are used?

☐Paracetamol

☐Ibuprofen

☐Diclofenac

☐Ketorolac

☐Dipyrone

☐Other NSAIDs

☐Tramadol

☐Morphine

☐Fentanyl

☐Other opioids

☐Other

How frequently is analgesia done for politrauma associated pain?

☐Always

☐> 50% of the times

☐< 50% of the times

☐Never

Which drugs are used?

☐Paracetamol

☐Ibuprofen

☐Diclofenac

☐Ketorolac

☐Dipyrone

☐Other NSAIDs

☐Tramadol

☐Morphine

☐Fentanyl

☐Other opioids

☐Other

How frequently is analgesia done for burn associated pain?

☐Always

☐> 50% of the times

☐< 50% of the times

☐Never

Which drugs are used?

☐Paracetamol

☐Ibuprofen

☐Diclofenac

☐Ketorolac

☐Dipyrone

☐Other NSAIDs

☐Tramadol

☐Morphine

☐Fentanyl

☐Other opioids

☐Other

Procedural Analgesia and Sedation

Is sucrose used for painful procedures in infants <6 months years old?

☐Yes

☐No

Is EMONO used for procedural sedoanalgesia?

☐Yes

☐No

Is EMLA® used for procedural analgesia?

☐Yes

☐No

Is local analgesia done for intramuscular injections?

☐Yes

☐No

How frequently is analgesia done for venopuncture?

☐Always

☐> 50% of the times

☐< 50% of the times

☐Never

What kind of analgesia is used in these situations?

________________________________________________________________

How frequently is analgesia done for lumbar puncture?

☐Always

☐> 50% of the times

☐< 50% of the times

☐Never

What kind of analgesia is used in these situations?

________________________________________________________________

How frequently is sedation done for lumbar puncture?

☐Always

☐> 50% of the times

☐< 50% of the times

☐Never

Which drugs do you use for lumbar puncture sedations?

________________________________________________________________

How frequently is analgesia done for fracture reductions?

☐Always

☐> 50% of the times

☐< 50% of the times

☐Never

What kind of analgesia is used in these situations?

________________________________________________________________

How frequently is sedation done for fracture reductions?

☐Always

☐> 50% of the times

☐< 50% of the times

☐Never

Which drugs do you use for these sedations?

________________________________________________________________

Where are fracture reductions done?

☐Orthopedic emergency department

☐PED

☐Other

How frequently is analgesia done for wound disinfection and suture?

☐Always

☐> 50% of the times

☐< 50% of the times

☐Never

What kind of analgesia is used in these situations?

________________________________________________________________

How frequently is sedation done for wound disinfection and suture?

☐Always

☐> 50% of the times

☐< 50% of the times

☐Never

Which drugs do you use for these sedations?

________________________________________________________________

Where are wound disinfections and sutures done?

☐Surgery emergency department

☐PED

☐Other

Who is responsible for the IV procedural sedoanalgesia?

☐Pediatrician

☐Anesthetists

☐Any physician

Is there support for intensivists or anesthetists moderate to profound sedation?

☐Yes

☐No

Is pre-sedation risk evaluated?

☐Yes

☐No

Is pre-sedation risk, monitoring and drugs used registered after the procedure?

☐Yes

☐No

General Use of Opioids

Opioid prescription is done by:

☐Any physician of the PED

☐Only pediatricians

☐Only with the intensivists or anesthetists support

☐Other

Non-Pharmacological Interventions

Are non-pharmacological interventions used?

☐Yes

☐No

How frequently is forced immobilization used?

☐Always

☐> 50% of the times

☐< 50% of the times

☐Never

How frequently are parents present for procedures?

☐Always

☐> 50% of the times

☐< 50% of the times

☐Never

How frequently is the parent’s lap used?

☐Always

☐> 50% of the times

☐< 50% of the times

☐Never

Training and Adequacy of Pain Treatment

Do you think that pain is adequately managed at your PED?

☐Yes

☐No

If you do not think so, why?

☐Difficulty in communicating with children

☐ Difficulty of a correct pain assessment

☐Inadequate methods

☐Lack of time

☐Lack of motivation

☐Lack of material

☐Inadequate medical prescriptions

☐Medical inexperience in prescribing analgesics

☐Fear of hiding symptoms and signs

☐Fear of opioid side effects

☐Insufficient child and parents’ information

☐Insufficient staff knowledge of how to manage pain

☐Procedures are too quick, they don’t need analgesia

☐Pain is natural, we should not treat it every time

☐Other

If you responded “other”, please specify it

___________________________________________________________________

Do you think that your staff should have more training of in pain management?

☐Yes

☐No

Comments

___________________________________________________________________
